# Dysautonomia in long COVID is prevalent and could explain the frequency of symptoms

**DOI:** 10.1101/2025.03.24.25324564

**Authors:** Leonardo Tamariz, Irina Rozenfeld, Rafael Iglesias, Elizabeth Bast, Santiago Avecillas, Lina Shehadeh, Nancy Klimas, Ana Palacio

## Abstract

**Background:** Long COVID presents with a variety of symptoms. Some of those symptoms could be related to autonomic dysfunction. Our aim is to evaluate the prevalence of autonomic dysfunction in long COVID patients.

**Methods:** We conducted a cross-sectional study and included all consecutive patients enrolled in several clinical research studies. We performed the following autonomic dysfunction markers: heart rate variability, heart rate, systolic and diastolic blood pressure changes during NASA lean test, cardiopulmonary exercise testing and a COMPASS-31 scale. We used linear regression to calculate the contribution of each dysautonomia measure on symptom burden as measured by the modified COVID-19 Yorkshire scale.

**Results:** We included 100 patients for this study. Our sample had a mean age of 56+/-11 years, included 53% minorities and 32% were women. Dysautonomia as defined by an abnormal COMPASS-31 was seen in 82%; 95% 72-89 while cardiovascular resting dysautonomia as represented by an abnormal heart rate variability was seen in 60%; 95% 48-70 of the population, orthostatic hypotension in 12% and POTS in 10%. In our adjusted analysis, we found that the beta coefficient for the COMPASS-31 score (0.37) was significant on changes in a self-reported long COVID symptom burden. The orthostatic intolerance and gastrointestinal domains of the COMPASS-31 was associated the highest long COVID symptom burden.

**Conclusion:** Dysautonomia is common in long COVID patients and contributes to the overall symptoms seen in long COVID. Identifying dysautonomia has important diagnostic and therapeutic implications.

## Background

Long COVID is a complex disease that presents with a variety of symptoms.[1] The prevalence of long COVID in the United States ranges from 7-19%. [2][3] The growing research in the area has already identified several mechanisms like endothelial dysfunction, viral reactivation, mast cell activation, immune exhaustion and autonomic dysfunction as prevalent and potentially responsible for the symptomatology.[1]

Among those, dysfunction of the autonomic nervous system (ANS) is emerging as one of the most prevalent abnormalities.[4] The role of the ANS in a condition that affects most organ systems would not be surprising since the ANS innervates all organs and is responsible for numerous physiological functions. Other conditions with multi-system symptoms such as myalgic encephalomyelitis / chronic fatigue syndrome (ME/CFS) and fibromyalgia have reported autonomic dysfunction as a mediator of those conditions. [5] Yet, the mechanism(s) are not fully elucidated. In long COVID there is evidence of immune system dysfunction, mast cell activation and a chronic stress response that leads to a persistent state of flight or fight.[6] However, it is unclear if these mechanisms are caused by the direct effect of the virus on the brainstem or through endothelial cell damage, versus an indirect mechanism mediated by the persistent activation of the immune system. [1]The possibility that all these mechanisms co-exist and mediate many of the abnormalities found, is non-trivial and should be the focus of phenotyping research studies.

For clinical purposes though, it is urgent to evaluate and report the best strategies to identify autonomic dysfunction that affects the long COVID symptom burden. Having accurate measures would allow providers to understand what organ-systems are experiencing the most dysregulation, the potential mechanisms of the symptomatology and guide the treatment. The existing published recommendations recommend evaluating orthostatic intolerance, heart rate variability and self-reported global dysautonomia using a questionnaire such as the COMPASS-31.[7] The purpose of this study was to evaluate which of these measures best correlates with long COVID symptom burden and are more likely to offer therapeutic insights to providers.

## Methods

### Study setting

The study collected information from two sites. The Veteran Affairs healthcare system (VAHS) and the Institute for NeuroImmune Medicine (INIM) from the NOVA Southeastern University. The Miami VAHS long COVID clinic was established in 2021 as a multidisciplinary clinic that offers hybrid services (virtual or in person). Patients are referred by primary care providers or screened via a national VAHS digital screening pilot. The INIM at NOVA was established in 2014 as a multidisciplinary clinic that offers in person care for patients with ME/CFS, fibromyalgia and post-COVID. The Miami VHAS and INIM clinics serve a racially diverse population in South Florida.

### Study design and study population

We conducted a cross-sectional study of consecutive non-selected patients evaluated at the two participating clinics and recruited from three ongoing long COVID research studies.

First, a pilot randomized clinical trial evaluating a behavioral/coaching intervention; second, a cross-sectional phenotyping study evaluating endothelial dysfunction, and third, an observational pilot practice-based research network (PBRN) study to evaluate preference and effectiveness of long COVID treatments among patients enrolled in a long COVID clinic. The inclusion criteria for all studies included a diagnosis of long COVID based on the WHO criteria and the presence of post COVID symptoms in the modified COVID-19 Yorkshire Rehabilitation Scale (C19-YRSm). Table 1 of the appendix provides relevant information and contribution from each study to this sample. The three studies used the same validated instruments to evaluate long COVID disease severity. For this study we only report the baseline assessments for all participants at study entry. All studies were approved by the research ethics committees at the Miami VAHS and NOVA Southeastern University and all participants signed informed consent prior to participation. Recruitment started depending on the study for the PBRN on November 1, 2024, for the phenotyping study on November 1, 2023 and the behavioral coaching intervention on June 1, 2023.

**Table 1:** Baseline characteristics of 100 long COVID patients.

|  |  |
| --- | --- |
| Number | 100 |
| Age, mean and SD | 56.6+/-11 |
| Female, % | 32% |
| Black, % | 26% |
| Hispanic, % | 27% |
| ME/CFS, % | 23% |
| Diabetes, % | 16% |
| Pre-diabetes, % | 17% |
| Body mass index | 30.4+/-5.8 |
| Beta-blocker, % | 20% |
| Vagal nerve stimulator, % | 13% |

### Study Procedures

Once individuals were identified and consented for one of the trials, we scheduled them to complete an in-person baseline assessment that included multiple previously validated methodologies.

### Autonomic dysfunction testing and definition of dysautonomia

We evaluated dysautonomia using the following metrics: heart rate variability (HRV), changes in blood pressure and heart rate during the NASA lean test, exercise induced dysautonomia during a cardio-pulmonary exercise test and the COMPASS-31 score as a measure of self-reported global dysautonomia.

*Heart Rate variability (HRV):* We used the Heartmath device emWave Pro Plus to measure HRV. We used the HRV feature without deep breathing assessment to capture the frequency domains over 2 minutes. The validity of the short time period measures has been established.[8] We report the standard deviation of normal to normal (NN) intervals (SDNN). We defined HRV as abnormal if SDNN values were below 50 ms and borderline if SDNN values were between 50 and 100 ms.[8]

*Composite Autonomic Symptom Score 31 (COMPASS-31) questionnaire:* The COMPASS-31 is validated and widely used questionnaire to quantify autonomic symptom severity. It consists of 31 questions that fall into six domains of dysautonomia: orthostatic intolerance, vasomotor, secretomotor, gastrointestinal, bladder, and pupillomotor. An answer was scored as zero when it was not assigned a point. A raw domain score was obtained by adding together points within each domain. The total score within each domain was weighted and then added together to give a total score ranging from 0 to 100. [9] A total COMPASS-31 of >28.6 was used to suggest initial autonomic nervous system dysfunction, as reported in earlier studies.[10]

*NASA lean test:* We used the NASA lean test to evaluate the effect of standing on blood pressure and heart rate. The test is validated and has accurate changes when compared to a tilt test.[11] During the test the participant has his/her blood pressure and pulse measured lying down, then each minute for 10 minutes while standing leaning against the wall. We defined orthostatic hypotension as a difference of ≥20 mmHg in the systolic blood pressure and/or ≥10 mmHg in the diastolic blood pressure between each baseline value and its minimum reading between minutes 1, 2, and 3 and postural tachycardia as an increase >30 bpm or to >120 bpm within 10 minutes of position change in the absence of orthostatic hypotension. [9] In addition, we report the largest difference in systolic blood pressure and heart rate and from supine to standing that occurs over the course of the test.

*Cardiopulmonary Exercise Testing (CPET):* We evaluate for exercise induced autonomic dysfunction in selected patients who received CPET as a part of their clinical care [12] CPET was performed on an electromagnetically braked cycle ergometer at 16° to 18°C, with indirect calorimetry. The CPET protocol followed at least 5 minutes of rest. Two-minute resting recording was followed by 2-minute unloaded cycling at a cadence of 60 rpm, before introduction of a 25-W resistance followed by a progressively increasing workload to volitional fatigue. Here we report only exercise induced dysautonomia defined as the Jouven’s criteria: (1) resting HR >75 bpm; (2) increase in HR during exercise <89 bpm; and (3) HR recovery <25 bpm in the first 60 s after cessation of exercise. [13]

### Long COVID symptoms

We evaluated long COVID symptoms using the modified COVID-19 Yorkshire Rehabilitation scale (C19-YRSm). The C19-CYRSm is a 17-item instrument with subscales (scores): symptom severity (0–30), functional disability (0–15), other symptoms (0–25), and overall health (0–10). The scale has been validated in long COVID and the scale content covers all aspects of the WHO ICF framework. For the purpose of this study we used the symptom severity score as the burden of symptoms in long COVID.[14]

### Covariates

We collected demographic and clinical characteristics to include in our analysis. From the electronic health record (EHR) we collected age, gender and race/ethnicity defined as Non-Hispanic White, Non-Hispanic Black or Hispanic. We also collected clinical predictors known to be associated with dysautonomia: body mass index, the diagnoses of diabetes or prediabetes, hypertension, or ME/CFS as well as related treatments such as beta blockers and use of vagal nerve stimulators.

### Statistical analysis

We report baseline characteristics as mean with standard deviation and percentages.

We calculated the prevalence of each of the dysautonomia measures by dividing the number of patients with an abnormal dysautonomia measure by the entire sample and calculated the 95% confidence interval of the proportion using the binomial exact method.

To identify the contribution of each of the dysautonomia measures on long COVID symptoms we used different statistical tests. First, in our descriptive analysis, we reported by tertile of the C19-CYRSm symptom severity score, the mean (SD) and frequency of each dysautonomia measure. Second, we used linear regression with CYRS-19 symptom severity score as dependent variable and each dysautonomia measure, including the domains of the COMPASS 31, as independent variables. We reported the beta-coefficient of the change of the independent variable and the R-squared of the model as the percentage of the variance that the independent variable (dysautonomia measure or component of a measure) explained the dependent variable. We adjusted all models for age, gender, body mass index and the presence of diabetes. Third, we calculated a receiver operating characteristic curve (ROC). To calculate the ROC we categorized the dependent variable C19-YRSm symptom severity into severe (symptom score >20) and moderate (symptom score <20). We used each dysautonomia measure as the independent variable as the predictor variable and also combined the dysautonomia variables. When combining predictor variables, the highest category included two abnormal dysautonomia measures, the intermediate category had one abnormal variable and the lowest category had two normal dysautonomia variables.

The fitness of the data was assessed using the deviance ratio. Analyses were performed using STATA version 17 (College Station, Texas), and all significance tests were two-tailed.

## Results

### Baseline characteristics

We included 100 patients for this study. Table 1 shows the baseline characteristics of the 100 patients with long COVID enrolled across the three studies described. Our sample had a mean age of 56+/-11 years, included 53% minorities and 32% were women. Our long COVID patients had a high prevalence of a prior ME/CFS diagnosis (23%) and diabetes (16%) and had a mean BMI of 30.4+/-5.8. Thirteen percent of our patients used a GammaCore Sapphire transcutaneous vagal nerve stimulator prior to entering any of the studies and 20% used a beta-blocker. Table 2 of the appendix compares the C19-CYRSm symptom severity score for the entire long COVID clinic population and those who participated in the research studies revealing that those enrolled had a disease severity that was representative of the whole.

**Table 2:** Prevalence of dysautonomia measures by cut-offs.

| Number | Measure | Cut-off | Prevalence |
| --- | --- | --- | --- |
| 100 | SDNN | <100 | 94% |
|  |  | <50 | 60% |
| 100 | COMPASS | >28.7 | 82% |
| 100 | Orthostatic hypotension | >20 drop in SBP | 12% |
|  |  | >10 drop in DBP | 10% |
| 100 | POTS | >30 bpm | 10% |
| 25 | CPET-exercise induced autonomic dysfunction | Number of Jouven criteria |  |
|  |  | 3 | 57% |
|  |  | 2 | 100% |

### Prevalence of dysautonomia

Table 2 shows the prevalence of dysautonomia by measure used. Dysautonomia as defined by an abnormal COMPASS-31 was seen in 82%; 95% 72-89 while cardiovascular resting dysautonomia as represented by an abnormal heart rate variability was seen in 60%; 95% 48-70 of the population, orthostatic hypotension in 12% and POTS in 10%. Measures of exercise induced dysautonomia were seen in 57% among the 25 long COVID patients who had a CPET.

### Contribution of dysautonomia to long COVID symptoms

We found a dose effect in the impact of self-reported dysautonomia via the COMPASS-31 score and long COVID symptoms. Those in the highest tertile of C19-CYRSm symptom severity had a significantly higher (p=0.01) COMPASS-31 compared to those in the lowest quartile. (figure 1) We found a similar trend between blood pressure and heart rate changes during NASA Lean test and the tertiles of C19-CYRS, but results were not significant (p>0.05). In our adjusted analysis, we also found that the beta coefficient for the COMPASS-31 score was significant, reinforcing the finding that changes in a self-reported dysautonomia score correlate with long COVID symptom burden. We also found that the beta coefficient was significant for domains of the COMPASS-31 except for vasomotor suggesting that these particular domains are most strongly predictive of long COVID symptoms (table 4). Gastrointestinal and orthostatic intolerance were associated with highest long COVID symptoms.

**Figure 1:**
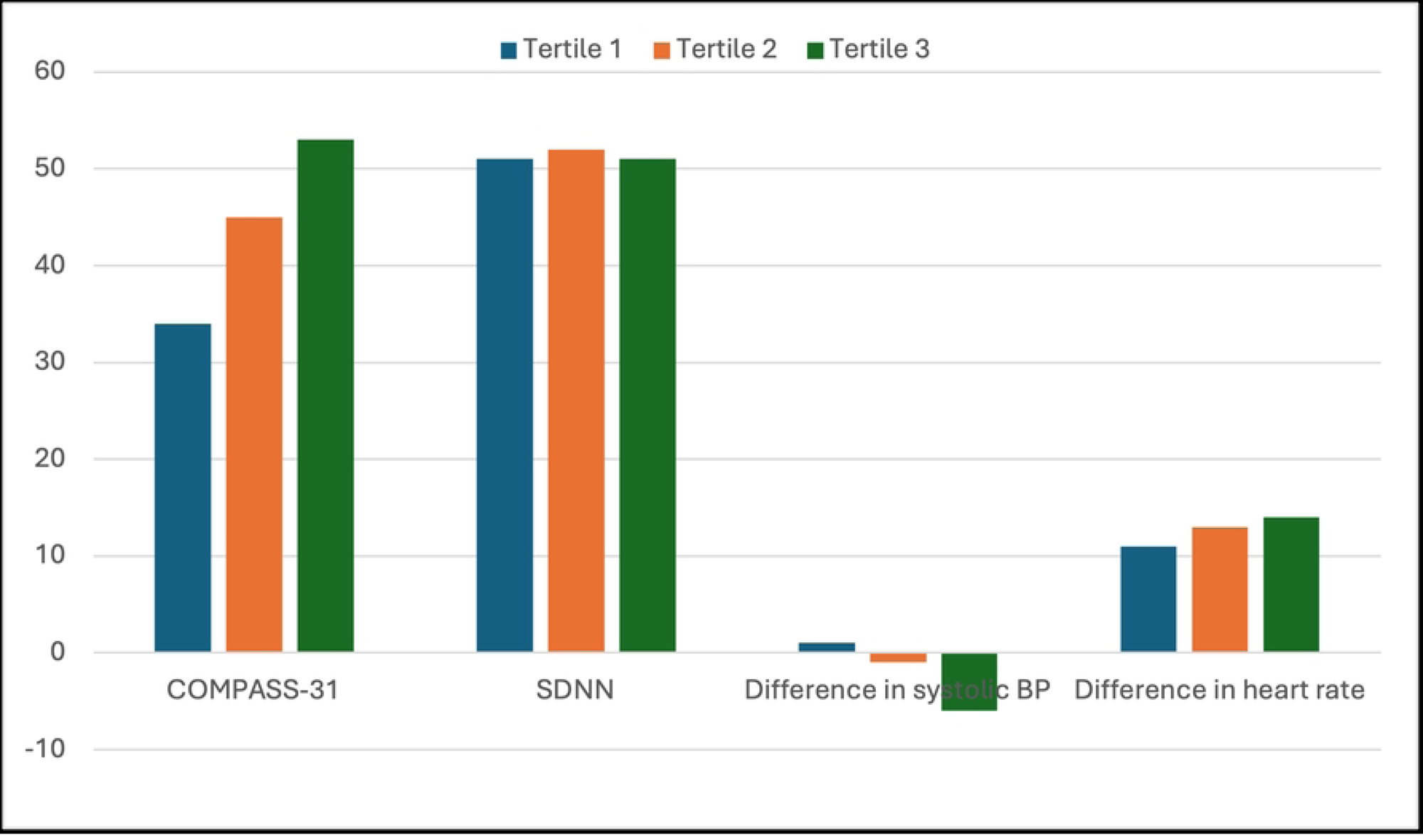
Distribution of the dysautonomia measures by tertile of C19-CYRSm

**Table 3:** Beta-coefficient by measure of dysautonomia on long COVID symptoms.

| Measure | Beta-coefficient (p-value) of increasing C19-YRSm | R-squared |
| --- | --- | --- |
| COMPASS-31 | 0.37 (<0.01) | 32% |
| Decreasing SDNN | -0.06 (0.46) | 7% |
| Decrease in systolic blood pressure during the NASA lean test | 0.12 (0.54) | 10% |
| Decrease in diastolic blood pressure during the NASA lean test | 0.08 (0.06) | 10% |
| Increase in heart rate during the NASA lean test | 0.12 (0.05) | 14% |
| Jouven criteria | 6.5 (0.55) | 6% |

Adjusted for age, gender, ME/CFS, diabetes and BMI.

**Table 4:** Beta-coefficient by each component of the COMPASS-31 score on long COVID symptoms.

| Domains of the COMPASS-31 | Beta-coefficient (p-value) | R-square |
| --- | --- | --- |
| Orthostatic intolerance | 0.55 (0.01) | 22% |
| Vasomotor | 1.93 (0.21) | 12% |
| Secretomotor | 1.27 (0.01) | 21% |
| Gastrointestinal | 0.97 (<0.01) | 25% |
| Bladder | 1.79 (0.04) | 17% |
| Pupillomotor | 2.91 (0.04) | 17% |

Adjusted for age, gender, ME/CFS, diabetes and BMI.

The other dysautonomia measures had non-significant beta-coefficients. Evaluating the R squared of each independent variable again revealed that the COMPASS-31 explained the most variance of all the measures. (table 3). Nevertheless, the ROC calculations suggests that in addition to the COMPASS-31 score, a change in HR during the NASA lean test or a having exercise induced dysautonomia also has a significant area under the curve and that the addition of HR to the COMPASS-31 provides the highest ROC. (Appendix, table 4).

## Discussion

Our study confirms that dysregulation of the ANS is highly prevalent among patients diagnosed with long COVID, although the positivity of the various measures described varies. Self-reported global dysautonomia using a validated questionnaire (COMPASS-31), was more common and significantly correlated with the long COVID symptom burden when compared to objective cardiovascular measures of dysautonomia, this is consistent with previous studies. We also found that specific domains of the COMPASS-31 were associated with long COVID symptom burden. The strength of our study includes the use of validated and standardized assessments for other clinical research studies and the consistency of the results across multiple statistical tests.

There are several limitations that deserve mention. First, we included participants who had agreed to participate in our research studies, therefore selection bias could have occurred. Nevertheless, the participating sample is representative of our clinics as demonstrated by a comparable C-19 YRSm symptom severity score. Second, the COMPASS-31 is a subjective measure with many non-specific symptoms that may overlap with other conditions (e.g. dizziness, diarrhea and/or constipation, urinary issues), which may be difficult to interpret in the absence of known dysautonomia. Second, our sample size was not large enough to allow for more covariates in the adjusted model and to potentially find statistical significance of certain metrics. Third, not all patients had a CPET and therefore our non-significant results for exercise-induced autonomic dysfunction could be explained by the small sample. Fourth, we did not have a control group and therefore are unable to compare dysautonomia results directly with non-long COVID patients, however the COMPASS-31 has previously been used in samples with healthy controls. Finally, the objective measures, were collected at one point in time which may decrease our ability to find associations, since these variables can be dynamically influenced by several factors daily.

There is strong evidence that long COVID is associated with dysregulation of the ANS. [9][15] There is considerable attention being given to cardiovascular metrics of dysautonomia.[16][17] Recently, cardiovascular dysautonomia, defined as POTS or orthostatic hypotension, was identified as an important component of post COVID syndrome and this finding has been associated with microvascular dysfunction leading to tissue ischemia. A study using the COMPASS-31 survey done globally in 2,413 adults found that 66% of all long COVID patients had moderate to severe autonomic dysfunction (score >20) independent of hospitalization for COVID-19. [18]We found that almost all our patients had a COMPASS-31 score greater than 20 and this could be explained by the fact that our patients were seen in a long COVID clinic while in the study by Larsen et al. participants came from support groups. A multidisciplinary collaborative consensus recommended several tests to evaluate dysautonomia and treatment for this group of patients but did not recommend the use of the COMPASS-31.[7] There is no controversy that finding an objective measure of dysautonomia, such as a significant change in blood pressure or heart rate when standing, is confirmatory and can be extremely helpful to guide treatment. However, in our clinic we discovered that only 25% of patients have an abnormal finding during a clinic visit. This may be related to the fact that, as many patients report to us, there are better days than others, and the objective metrics are dynamic and can be influenced by temperature, food and fluid intake, amount of effort done in the day, stress, amount of sleep, etc. Novak et al previously reported a mismatch between objective and subjective tests for dysautonomia which suggests that cross-sectional assessments may not be sufficient to assess ANS function. [19]In contrast, the self-reported questionnaire asks about symptoms over a period and thus may be more reflective of the overall patient experience. The COMPASS-31 also assesses multiple domains of autonomic dysfunction while current objective measures focus on cardiovascular dysautonomia which may under count other manifestations. The COMPASS-31 survey is a useful tool that can evaluate dysautonomia in different organ systems and these symptoms associated significantly with long COVID symptoms.

An alternative explanation is that the various dysautonomia measures reflect a different pathophysiology. The etiology of dysautonomia in long COVID is multifactorial and not completely understood. Theories include direct viral damage of the nerves, autoantibodies and inflammatory mechanisms, and microvasculature dysfunction. A study by Woo et al. performed histopathological characterization of postmortem vagus nerves from COVID-19 patients and controls, and detected SARS-CoV-2 RNA together with inflammatory cell infiltration.[20] A study by El-Rhermoul et al. reviewed the association between POTS and autoantibodies: auto-antibodies to the α and β receptors on GPCR antibodies, ganglionic acetylcholine receptor antibodies, angiotensin II receptor antibodies and antibodies to structural cardiac proteins have been identified in POTS. [21]The presence of autoantibodies can correlate with symptoms such as gastrointestinal disturbance, fatigue, muscle pain, exercise tolerance and standing time. Another theory includes the endothelial dysfunction and microvascular dysregulation due to over sympathetic drive. [22]

Our study highlights the need for cardiovascular (NASA lean test or tile table test, HRV, CPET) and non-cardiovascular (COMPASS-31) dysautonomia evaluation. Finding POTS or orthostatic hypotension in a patient can guide treatment: there are well established non-pharmacologic measure such as compression, salt and electrolytes, as well as use of beta blockers and or ivabradine that may be considered. Finding an abnormal HRV can help in screening future cardiovascular complications if other cardiovascular risk factors are present. However, lack of objective measures does not rule out dysautonomia in long COVID patients. The COMPASS-31 is a good screening tool that can prompt a closer follow up of objective measures. In addition, knowing a patient’s COMPASS score is particularly useful because it can identify specific organ that need to be targeted.

Another important finding of our study was the contribution of specific domains of the COMPASS-31 to long COVID symptoms. We found that all domains except for the vasomotor domain were associated with long COVID symptoms. Gastrointestinal and orthostatic intolerance were the most important contributors of symptoms. An interesting finding was the fact that patients reported orthostatic intolerance and did not meet the criteria for POTS or orthostatic hypotension. This requires further evaluation with a tilt test but potential explanations for this finding include the use of beta-blockers or the lack of validity of self-reported orthostatic intolerance.

In conclusion, dysautonomia is prevalent in long COVID, using different instruments can identify cardiovascular or non-cardiovascular manifestations of dysautonomia and can trigger different evaluations and treatments.

## Appendix

**Table 1:** Aims, funding and contribution of each study to the sample.

| Study | Start date | Aims | Number of participants included | Funding |
| --- | --- | --- | --- | --- |
| Behavioral coaching intervention | November 2022 | Evaluate the effect of a behavioral intervention on long COVID symptoms and measures of autonomic dysfunction | 45 | NOVA Southeastern University |
| Deep phenotyping study | November 2023 | Evaluate blood biomarkers and the vascular reactivity index in patients with long COVID and controls | 45 | American Heart Association |
| Practice based research network | November 2024 | Evaluate combinations of treatments for long COVID | 10 | Veterans Affairs HSRD |

**Table 2:**
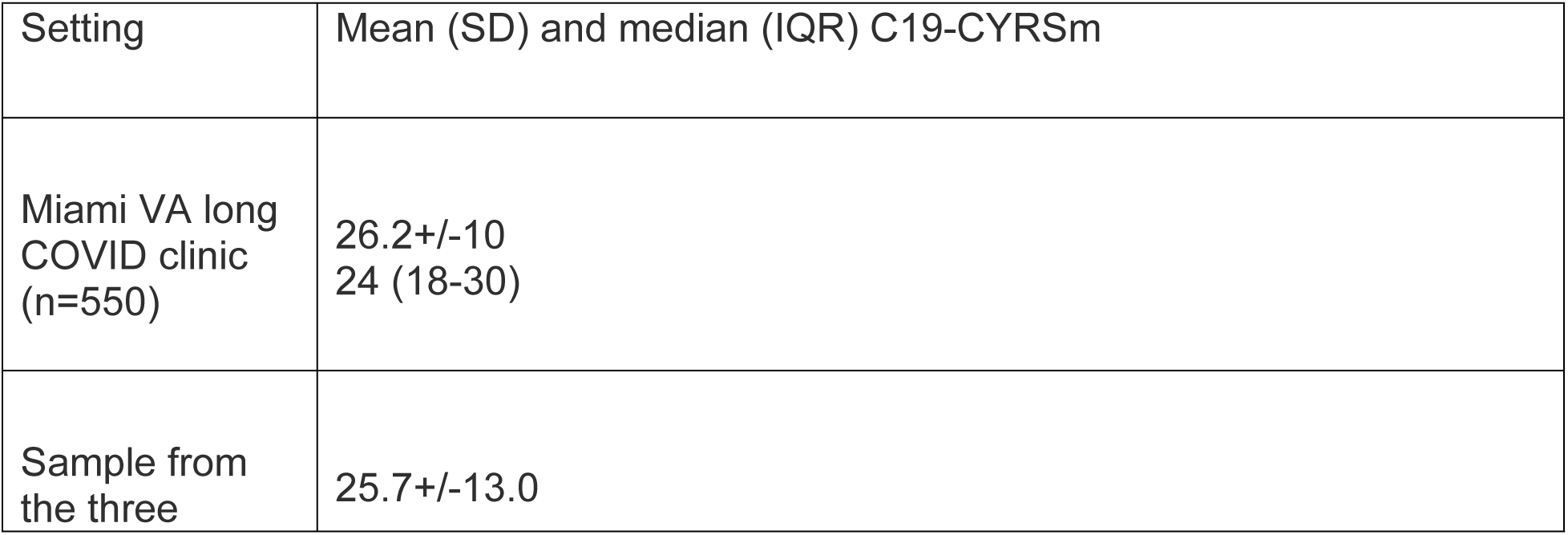
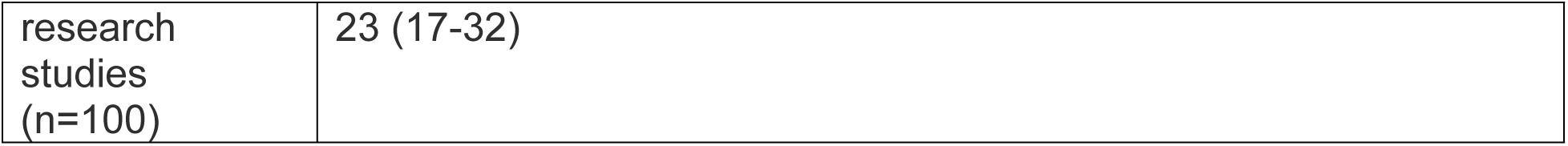
Miami VA Clinic and sample C19-YRSm.

**Table 3:**
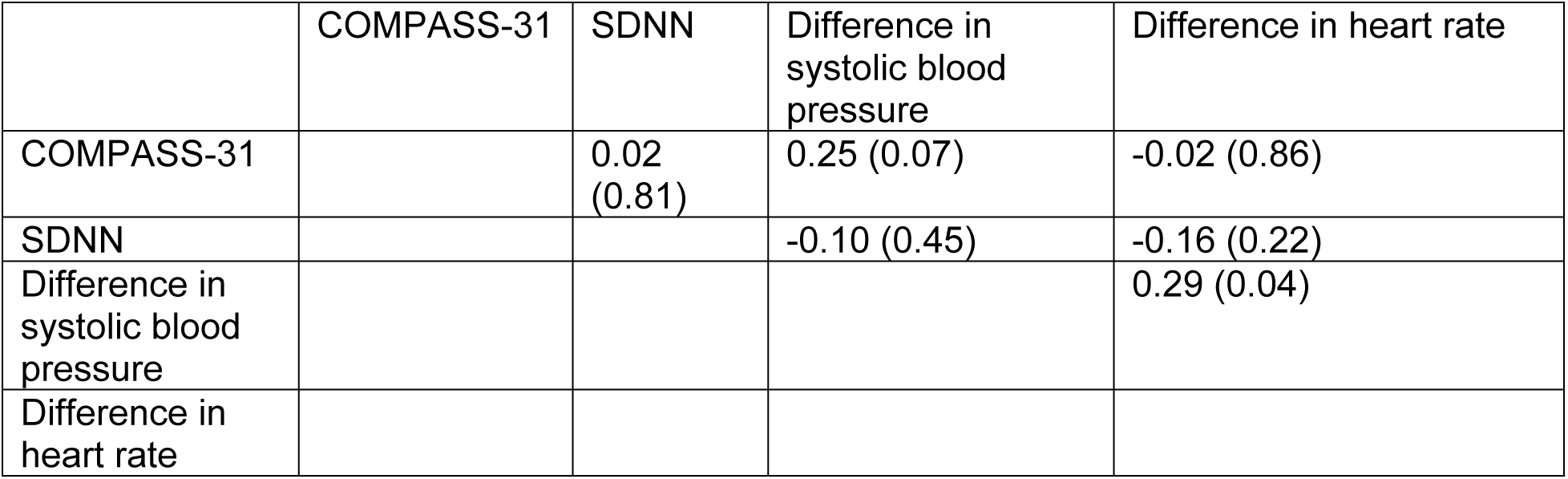
Correlation matrix of objective and subjective autonomic measures.

|  | COMPASS-31 | SDNN | Difference in systolic blood pressure | Difference in heart rate |
| --- | --- | --- | --- | --- |
| COMPASS-31 |  | 0.02 (0.81) | 0.25 (0.07) | -0.02 (0.86) |
| SDNN |  |  | -0.10 (0.45) | -0.16 (0.22) |
| Difference in systolic blood pressure |  |  |  | 0.29 (0.04) |
| Difference in heart rate |  |  |  |  |

**Table 4:** Areas under the ROC curve for each dysautonomia and combinations.

| Measure | Area under the ROC curve (95% CI) |
| --- | --- |
| Increasing SDNN | 57 (41-73) |
| COMPASS-31 | 69 (57-76) |
| Decrease in systolic blood pressure during the NASA lean test | 55 (36-73) |
| Decrease in diastolic blood pressure during the NASA lean test | 46 (27-65) |
| Increase in heart rate during the NASA lean test | 64 (55-79) |
| Jouven criteria | 68 (46-89) |
| COMPASS-31 + SDNN | 61 (51-64) |
| COMPASS-31 + changes in heart rate during the NASA lean test | 72 (61-81) |
| SDNN+ changes in heart rate during the NASA lean test | 53 (42-65) |

## Data Availability

Data cannot be shared publicly because it is owned but the Veterans Affairs.

